# Who do not wash their hands during the Covid-19 pandemic? Social media use as a potential predictor

**DOI:** 10.1101/2020.06.01.20119230

**Authors:** Stephen X. Zhang, Lorenz Graf-Vlachy, Rui Su, Jizhen Li, Kim Hoe Looi

## Abstract

This study predicts handwashing behavior during the Covid-19 pandemic. An analysis of 674 adults in Malaysia identifies their time spent on social media per day as a key predictor of handwashing. The association between time spent on social media and handwashing substantially depends on gender and the number of children in the same household. Additional predictors include age and health condition. This study helps identify specific target groups for health communication on hand hygiene via people’s use of social media, which can be a key channel for health communication campaigns during a pandemic.

## 1. INTRODUCTION

Handwashing is one of the most basic and effective measures to prevent the spread of disease in general [1] and of upper respiratory infections in particular.[2] Handwashing is notably recommended during pandemics.[3,4] Unfortunately, compliance remains a constant issue even among highly sophisticated populations like physicians and nurses.[5,6]

Given that health communication, such as messaging and reminders, has positive effects on handwashing behavior [7,8], it is advisable to identify, inform, and prompt at-risk populations to regularly wash hands during a pandemic. To economize on limited resources while having optimum impact, health organizations, governments, and other parties may want to focus their health messaging on less compliant groups.

This paper aims to identify predictors of handwashing behavior to help identify such less compliant groups. Specifically, we examine time spent on social media, which is a key channel for health communication during pandemics.[9] Social media has the advantage that at-risk populations, once identified, can directly be targeted with health information via the same platform.

However, to our best knowledge, there exist no studies connecting social media use to hand washing in any epidemic. This study identifies social media use and several demographic variables as predictors that may be useful when deploying targeted health information campaigns on hand hygiene during the ongoing Covid-19 crisis and future pandemics.

## 2. METHODS

### 2.1 Sample and data collection

We conducted a survey of working adults in Malaysia between May 2–8, 2020, six weeks after Malaysia implemented a *cordon sanitaire* preventive measure to contain COVID-19. To collect a reasonably representative sample in Malaysia, a multi-lingual and multi-ethnic country, the English version of the questionnaire was translated into Malay and Mandarin, the major languages in Malaysia. Links to the survey in Malay, Mandarin, and English were distributed via WhatsApp, Facebook Messenger, and email, via two-stage stratified sampling in terms of ethnicity, gender, age, and geographical area.[10] The survey was approved by Tsinghua University (20200322), and all survey participants consented online before enrolling in the survey. The respondents could opt out at any time and were ensured confidentiality and anonymity. No personally identifying information was collected.

### 2.2 Measures

The demographic variables of interest in this study comprise of gender, number of children living in the same household, age, educational level, and overall health condition. [11,12] The behavioral variables of interest are social media usage (measured in hours per day) and frequency of handwashing after touching things outside the home [seven-point Likert scale: 1 = never; 2 = rarely (less than 10% of the time); 3 = occasionally (about 30% of the time); 4 = sometimes (about 50% of the time); 5 = frequently (about 70% of the time); 6 = usually (about 90% of the time); 7 = every time].

We used Stata 16.1 to perform an OLS regression on the unweighted data.

## 3. RESULTS

### 3.1 Descriptive results

We obtained 674 valid responses from adults across all Malaysian states and federal territories. 51.5% (347) respondents were female and 48.5% (327) male. Almost half of the respondents were living without children in the household (48%, 324), and progressively fewer people were living with an increasing number of children, e.g., 17.1% (115) indicated to be cohabiting with one child and only 0.6% (4) were living with more than five children.

Some 2.8% (19) of respondents indicated they never or rarely washed their hands after touching things outside the household. 45.5% (307) respondents indicated they usually or always washed their hands. 51.6% (348) indicated handwashing frequencies in between. Social media use was widespread in our sample. While 29.7% (100) of respondents reported less than two hours of social media use per day, 29.8% (201) reported between two and just below four hours, and 40.5% (273) reported four or more daily hours. Detailed descriptives for all predictors can be found in Table 1.

**Table 1.** Descriptive findings and predictors of handwashing (*n* = 674)

| Variables | Description | Coefficient (95% CI) |
| --- | --- | --- |
| Handwashing: Mean = 5.33, std. dev. = 1.40 |  |  |
| Gender (categorical) |  |  |
| Female | 347 (51.5%) | -0.25 (-0.72 to 0.22) |
| Male | 327 (48.5%) | Reference category |
| Age (continuous) |  |  |
| 20–29 | 103 (15.8%) | 0.03*** (0.02 to 0.04) |
| 30–39 | 198 (29.4%) |  |
| 40–49 | 193 (28.6%) |  |
| 50–59 | 148 (22.0%) |  |
| >59 | 32 (4.8%) |  |
| Educational level (continuous) |  |  |
| Primary school completed (1) | 0 (0.0%) | -0.04 (-0.23 to 0.14) |
| Secondary school completed (2) | 49 (7.3%) |  |
| College or university completed (3) | 410 (60.8%) |  |
| Graduate school completed (4) | 215 (31.9%) |  |
| Health condition (continuous) |  |  |
| Poor (1) | 7 (1.0%) | 0.17** (0.06 to 0.28) |
| Fair (2) | 74 (11.0%) |  |
| Good (3) | 212 (31.5%) |  |
| Very good (4) | 252 (37.4%) |  |
| Excellent (5) | 129 (19.1%) |  |
| Number of children in household (continuous) |  |  |
| 0 | 324 (48.0%) | -0.22* (-0.39 to -0.05) |
| 1 | 115 (17.1%) |  |
| 2 | 101 (15.0%) |  |
| 3 | 76 (11.3%) |  |
| 4 | 41 (6.1%) |  |
| 5 | 13 (1.9%) |  |
| > 5 | 4 (0.6%) |  |
| Time on social media per day (continuous) |  |  |
| [0h; 2h[ | 100 (29.7%) | -0.09** (-0.16 to -0.02) |
| [2h; 4h[ | 201 (29.8%) |  |
| [4h; 6h[ | 119 (17.7%) |  |
| [6h; 8h[ | 70 (10.4%) |  |
| > 8h | 84 (12.5%) |  |
| Interactions |  |  |
| Time spent on social media per day × Gender | - | 0.14** (0.06 to 0.23) |
| Time spent on social media per day × Number of children in household | - | 0.06** (0.03 to 0.09) |
| Gender × Number of children in household | - | 0.20 (-0.05 to 0.46) |
| Time spent on social media per day × Gender × Number of children in household | - | -0.06** (-0.11 to -0.02) |

Note: \* $p < 0.05$ ; \*\* $p < 0.01$ ; \*\*\* $p < 0.001$ .

### 3.2 Predictors of handwashing frequency

Table 1 shows the regression results for handwashing (F(10, 663) = 6.81, *p* = 0.000). Age (b = 0.03, 95% CI: 0.02 to 0.04, *p* = 0.000) and health condition (b = 0.17, 95% CI: 0.06 to 0.28, *p* = 0.003) were associated with frequency of handwashing, with older and healthier people washing their hands more frequently. Number of children (b = −0.22, 95% CI: −0.39 to −0.05, *p* = 0.010) and time spent on social media per day (b = −0.09, 95% CI: −0.16 to −0.02, *p* = 0.007) were significantly negatively related to handwashing.

The relationship between time spent on social media and handwashing, however, was significantly moderated by gender (b = 0.14, 95% CI: 0.06 to 0.23, *p* = 0.001) and number of children (b = 0.06, 95% CI: 0.03 to 0.09, *p* = 0.000). Further, there was a significant three-way interaction between time spent on social media, gender, and number of children (b = − 0.06, 95% CI: −0.11 to −0.02, *p* = 0.006). We next use margin analysis to break down and discuss the interaction results (see Figure 1).

**Figure 1.**
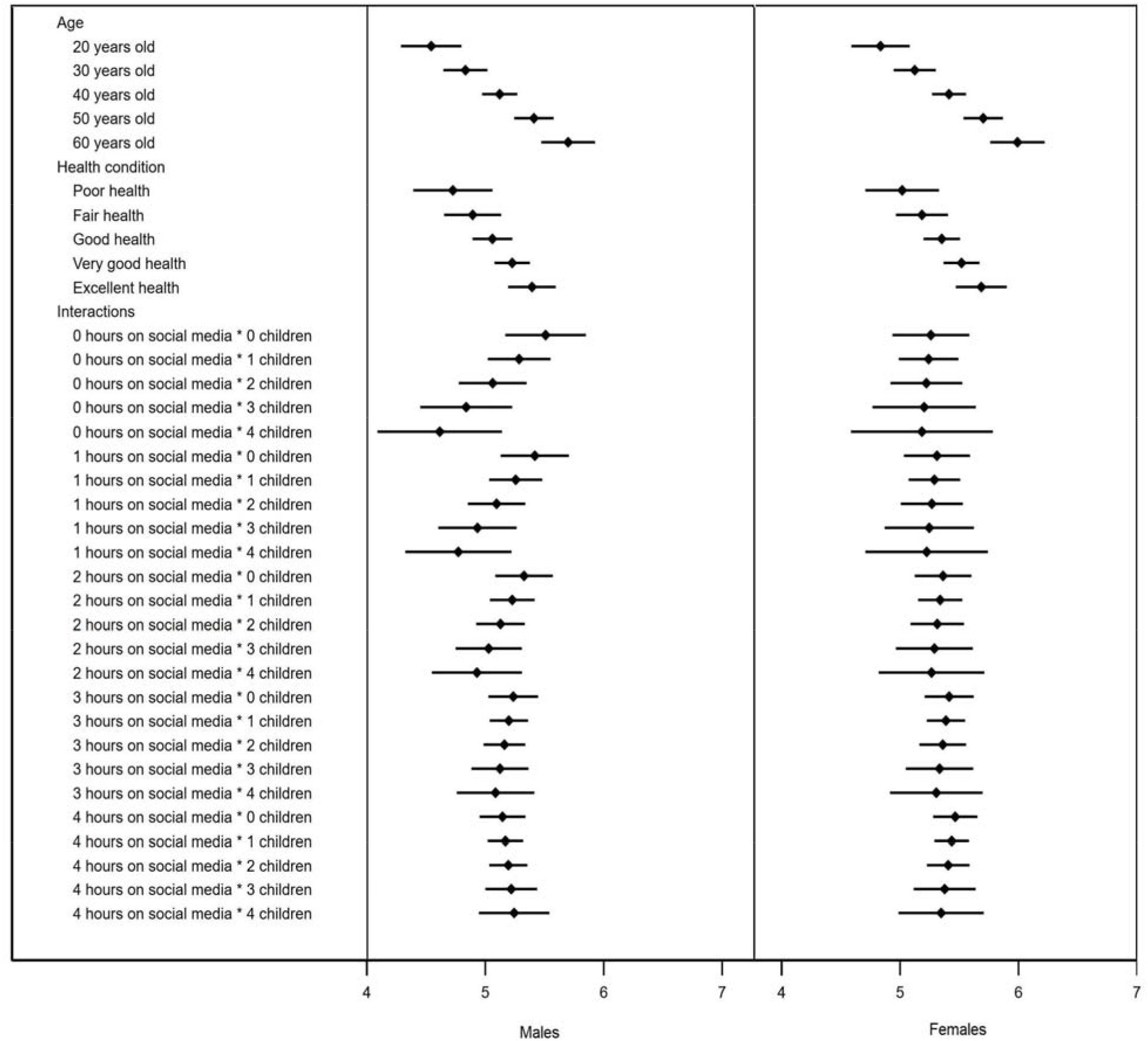
The predicted handwashing frequency at different values of the significant predictors (handwashing frequency @ 4 = sometimes (about 50% of the time); 5 = frequently (about 70% of the time); 6 = usually (about 90% of the time); 7 = every time)

## 4. DISCUSSION

Our results overall showed that social media use predicts handwashing, and that this predictor exhibits distinct patterns depending on gender and number of children. First, more time spent on social media was positively associated with the frequency of handwashing for males with three or more children living in the same household (e.g., for those with three children: b = 0.09, 95% CI: 0.02 to 0.17, *p* = 0.010). However, for males with no children in the same household, more time spent on social media was negatively associated with the frequency of handwashing (e.g., for those with no children: b = −0.09, 95% CI: −0.16 to −0.02, *p* = 0.007). The association between time spent on social media and handwashing was not significant for males with one or two children.

For females in general, the direct effect of social media use on handwashing was significant (b = 0.05, 95% CI: 0.005 to 0.09, *p* = 0.030). However, a margin analysis showed that more time spent on social media was significantly linked to more handwashing only for females with one child (b = 0.05, 95% CI: 0.005 to 0.09, *p* = 0.028) and marginally significant (p < 0.10) for females without children or with two children. The association between time spent on social media and handwashing was not significant for women with three or more children.

Second, prior studies frequently identified gender as a predictor of handwashing, finding that women generally wash their hands more than men both during pandemics and in other circumstances.[13, 14] While our results corroborate this direct effect of gender (b = 0.29, 95% CI: 0.08 to 0.50, *p* = 0.006), we also found that the gender difference depended on social media usage and the number of children. Specifically, our study revealed that there was no gender difference in handwashing between females and males who spent three hours or less on social media *(p* > 0.10 across all cases). The gender difference, i.e., the notion that females wash hands more than males do, was significant for those who spend more than three hours on social media and had no or one child (e.g., four hours social media and no children: b = 0.32, 95% CI: 0.05 to 0.59, *p* = 0.020). The gender difference was not significant for those with two or more children. Hence, the gender difference commonly found in the literature is not universal but instead depends on both social media use and the number of children. In sum, gender is a useful predictor of handwashing largely for people who spend lots of time on social media and have no or one child.

Third, this study also unveiled the number of children in the same household as an important predictor of hand washing. Number of children negatively predicted handwashing among males who did not use social media (b = −0.22, 95% CI: −0.39 to −0.05, *p* = 0.010) and who averaged one hour on social media (b = −0.16, 95% CI: −0.30 to −0.02, *p* = 0.028). In contrast, number of children positively predicted handwashing among males who spent lots of time social media (b = 0.21, 95% CI: 0.09 to 0.34, *p* = 0.001). The number of children did not predict handwashing for females in general.

Taken together, these results suggest that research on predictors of handwashing should not rely on gender, number of children, or social media use alone but must consider all three simultaneously to yield better predictions. This finding is increasingly relevant as social media is likely a key channel of information to people, and females and males tend to have different interests and social circles on social media [15], and people with varying numbers of children may similarly gravitate towards different interest groups. Thus, people receive social information – also regarding pandemics – differently depending on their gender and family situation. This study, as the first to identify social media use as a predictor of handwashing, suggests that it becomes increasingly critical to take social media usage into account in predictive models of human behavior, including in those of handwashing during a pandemic.

Finally, our results also provide evidence of the efficacy of other predictors of handwashing under the COVID-19 pandemic. Extant work found mixed evidence on the association between age and protective behaviors like handwashing during virus pandemics.[13] Our results suggest that there is indeed a positive relationship between age and handwashing. Prior work also found that more educated people tend to exhibit greater protective behavior during pandemics, but some results were inconclusive.[13] In our sample, there was no significant link between education and handwashing. We did, however, find a significant association between self-reported health condition and handwashing. During the pandemic, healthier people indicated greater handwashing frequency – a finding that is new to the literature. Hence, younger and less healthy people should generally be targeted.

Overall, our findings can be used to identify the population at risk to enable more targeted health communication. Reminders [16] and signage [8] about handwashing, and communication regarding a pandemic in general [7], are effective in increasing handwashing. Consequently, our research helps with the identification of the less compliant groups to enable more targeted delivery of such effective handwashing communications campaigns, especially via social media.

### 4.1 Limitations

First, the cross-sectional nature of our research precludes claims of causality, even though we are primarily interested in the predictive utility for screening less compliant groups. Second, self-reported compliance rates might be inflated due to social desirability [14], so our estimation of the less compliant groups is conservative. Third, we tried to get a representative sample via two-stage stratified sampling in terms of ethnicity, gender, age, and geographical area, but Malaysia is a multi-facet society and the sample should not strictly be considered nationally representative. Fourth, social media usage was very high during the COVID-19 pandemic in Malaysia due to a lockdown, and future studies may examine our model in lesser pandemics without strict lockdowns. Finally, Malaysia is an upper-middle income-level country, where water for handwashing is generally accessible. Studies in countries with heterogeneous individual access to water might yield different results.

### 4.2 Conclusion

We identify several new predictors of handwashing during a pandemic to enable health organizations, governments, and others to create more targeted communication campaigns, particularly via social media. Such campaigns might be particularly important because social media is likely used extensively during epidemics but most normal social media posts during a pandemic contain very little practical advice.[17] This study contributes by identifying less compliant groups regarding handwashing, especially via social media usage, to enable more focused health information campaigns.

## Data Availability

Will be available upon request once the paper is published.

## ACKNOWLEDGEMENT

We acknowledge the support of Tsinghua University-INDITEX Sustainable Development Fund (Project No. TISD201904).

